# Knowledge and misconceptions of the French population regarding medical genetics: a survey of 3,000 respondents

**DOI:** 10.64898/2026.07.17.26358259

**Authors:** Sandra Mercier, François Petit, Micheline Misrahi-Abadou, Philippe Berta, Anne Cambon-Thomsen, Boris Chaumette, Hervé Chneiweiss, Célia Crétolle, Patrick Edery, David Heard, Marina Konyukh, Chloé Laeng, Nizar Mahlaoui, Laurent Pasquier, Morgane Plutino, Sylvie Odent, Dominique Stoppa-Lyonnet, “Genetics and the General Public” FFGH Ethics Working Group

## Abstract

Advances in high-throughput sequencing and genetic research have expanded the role of genetics in medicine and society. Population-based screening programs, including neonatal and preconception testing, are increasingly implemented globally, alongside the rise of direct-to-consumer (DTC) genetic testing. The “Genetics and the General Public” Ethics Working Group of the French Federation of Human Genetics (FFGH) assessed knowledge and awareness of genetics within the French population through a nationally representative survey (n=3,013) conducted by the polling firm Ipsos bva. Results indicated that 69% of respondents report an interest in genetics, although their level of knowledge remains limited. Most respondents expressed positive attitudes toward genetics, perceiving it as a major source of hope in healthcare. While a majority indicated willingness to undergo genetic testing for medical purposes, they also reported legitimate concerns regarding the potential results. Despite legal restrictions, 12% reported having ordered a DTC genetic test (5% for genealogical; 5% for medical and 2% for both purposes), and 45% of non-users expressed strong interest in this type of test. Notably, there is a substantial lack of awareness regarding the limitations of these tests and the French legal framework governing their use. These findings highlight critical gaps in public knowledge, emphasizing the need for improved genetic education, including incorporating genetics into school curricula and launching targeted awareness campaigns. These initiatives should help clarify the distinctions between clinically validated genetic tests and DTC genetic testing services, addressing both their benefits and their ethical, legal, and scientific limitations, in order to promote informed decision-making.

## ARTICLE

Medical genetics is undergoing a profound transformation, driven by the increasing accessibility of high-throughput DNA sequencing technologies and the substantial reduction in associated costs in recent years.(1) In France, the France Médecine Génomique 2025 Plan enables whole-genome sequencing to be prescribed as a first-line test in specific clinical situations (referred to as ‘pre-indications’ in the Plan).(2) This evolution is accompanied by a continuous expansion of indications for molecular analyses, with medical genetics increasingly integrated across an increasing number of medical specialties.

In parallel, there is a growing trend towards implementing population-scale genetic testing strategies. For example, the PERIGENOMED research study, conducted within two French university hospital federations (FHU TRANSLAD and GenOMeDS), is investigating the feasibility and implications of expanding genomic newborn screening in France.(3) Internationally, approximately twenty research initiatives on genomic newborn screening are currently underway, with participating teams coordinated through the International Consortium on Newborn Sequencing.

Furthermore, several countries have introduced expanded carrier screening (ECS), including preconception screening for couples planning a pregnancy.(4) In France, prior to the 2018 revision of the bioethics law, the National Ethics Advisory Committee (Comité Consultatif National d’Ethique, CCNE) issued a favourable opinion on the development of preconception genetic screening provided that certain structural safeguards are observed, including a strictly voluntary and non-coercive approach.(5) However, this proposal was not adopted by the legislature. The issue is currently being revisited as part of the 2026 National Bioethics Conference to prepare for the upcoming revision of the Bioethics Act.

At the same time, despite being prohibited in France, direct-to-consumer (DTC) genetic testing is expanding through foreign online platforms. Although such testing is illegal under French law and subject to a €3,750 fine for individuals who order these tests, enforcement remains difficult in practice.

These developments raise important questions regarding autonomy in the genomic era. According to the principles articulated by Beauchamp and Childress, autonomy requires the capacity to act intentionally, with adequate understanding and free from controlling influences.(6) In France, prior free, informed, and written consent is legally required for any germline genetic testing. A key question, therefore, is whether the general population possesses the necessary knowledge to exercise such autonomy meaningfully. This issue is particularly relevant, as the responsible implementation of genomic medicine depends increasingly not only on technical capabilities, but also on the general public’s understanding, trust, and ability to distinguish between medically supervised testing and commercial genomic services. To address this issue, the Ethics Working Group “Genetics and the General Public” of the French Federation of Human Genetics (FFGH) conducted a national survey to assess public knowledge and misconceptions regarding genetics. The literature on public attitudes towards genetics remains limited, particularly in France.(7) To date, only one survey has explored public perceptions of genetic screening in the French population: a study conducted by Ipsos bva in 1988.(8) Given the substantial changes in the scientific, medical, and societal context since then, a new nationwide survey was conducted in November 2025 in collaboration with the polling firm Ipsos bva, involving 3,013 respondents representative of the French population. This manuscript presents the results of this survey and discusses perspectives for future action.

## SUBJECTS AND METHODS

### Survey design and data collection

The survey was developed in accordance with the objectives defined by ‘Genetics and the General Public’ working group of the FFGH, in collaboration with the polling firm Ipsos bva. The questionnaire comprised 18 items assessing knowledge of and attitudes towards genetics, as well as perceptions of medically indicated and direct-to-consumer genetic testing. See Supplementary Information (Document S2 : questionnaire in English and S3 in French).

Data collection was conducted by Ipsos bva using its proprietary online access, which includes approximately 350,000 individuals who have agreed to participate regularly in surveys via computer, smartphone or tablet. The panel is managed by Ipsos Interactive Services (IIS), a subsidiary specializing in online research.

In addition to quota variables, detailed sociodemographic data (including income, education level, employment characteristics and household composition) are available for panel members. Recruitment, engagement, and retention procedures comply with international quality standards to ensure data reliability. Panel members are recruited through multiple channels, including online advertising, offline media campaigns, telephone surveys, and referrals.

Panel entry involves reinforced verification procedures (updated in 2022), including detection of duplicate accounts, bots, and fraudulent profiles using technical controls such as IP address monitoring and device fingerprinting, as well as double email/SMS confirmation, and behavioral consistency controls. Following enrollment, panel members participate in surveys on a wide range of topics to minimize response bias and external influence.

Participants receive email invitations to complete the questionnaire developed jointly by the study group and Ipsos bva, without knowing in advance that the topic concerns medical genetics. The survey instrument was programmed and tested across multiple devices (smartphones and computers), followed by a pilot phase. Post-pilot quality checks included completion time, response consistency, dropout rates, quota monitoring, and device distribution. Fieldwork was continuously monitored to ensure appropriate targeting and sample representativeness. Data collection was fully anonymized, and the mean completion time was approximately 10 minutes.

Participants received compensation through a points-based system redeemable for vouchers or charitable donations. Incentives were modest and did not constitute direct financial remuneration.

Ipsos bva holds ISO 20252 (market research), ISO 27001 (information security), and ISO 9001 (quality management system) certifications.

### Sampling and statistical analysis

Quota sampling was performed based on the most recent national census data (INSEE), using the following variables: gender, age, socio-professional category, region and level of urbanization. The final sample is representative of the French population aged 18 years and older. Sociodemographic characteristics are summarized in Table 1.

**Table 1:**
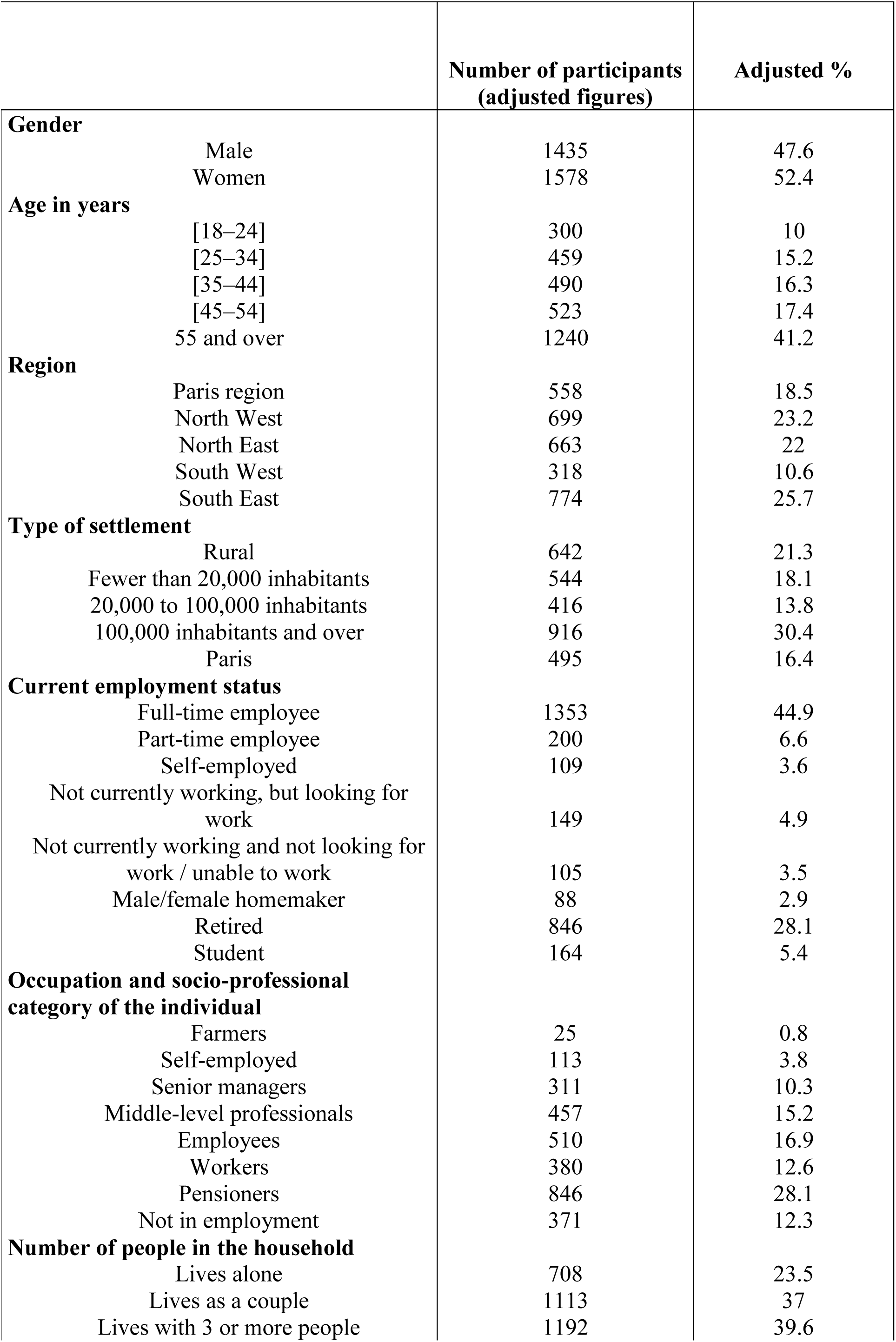
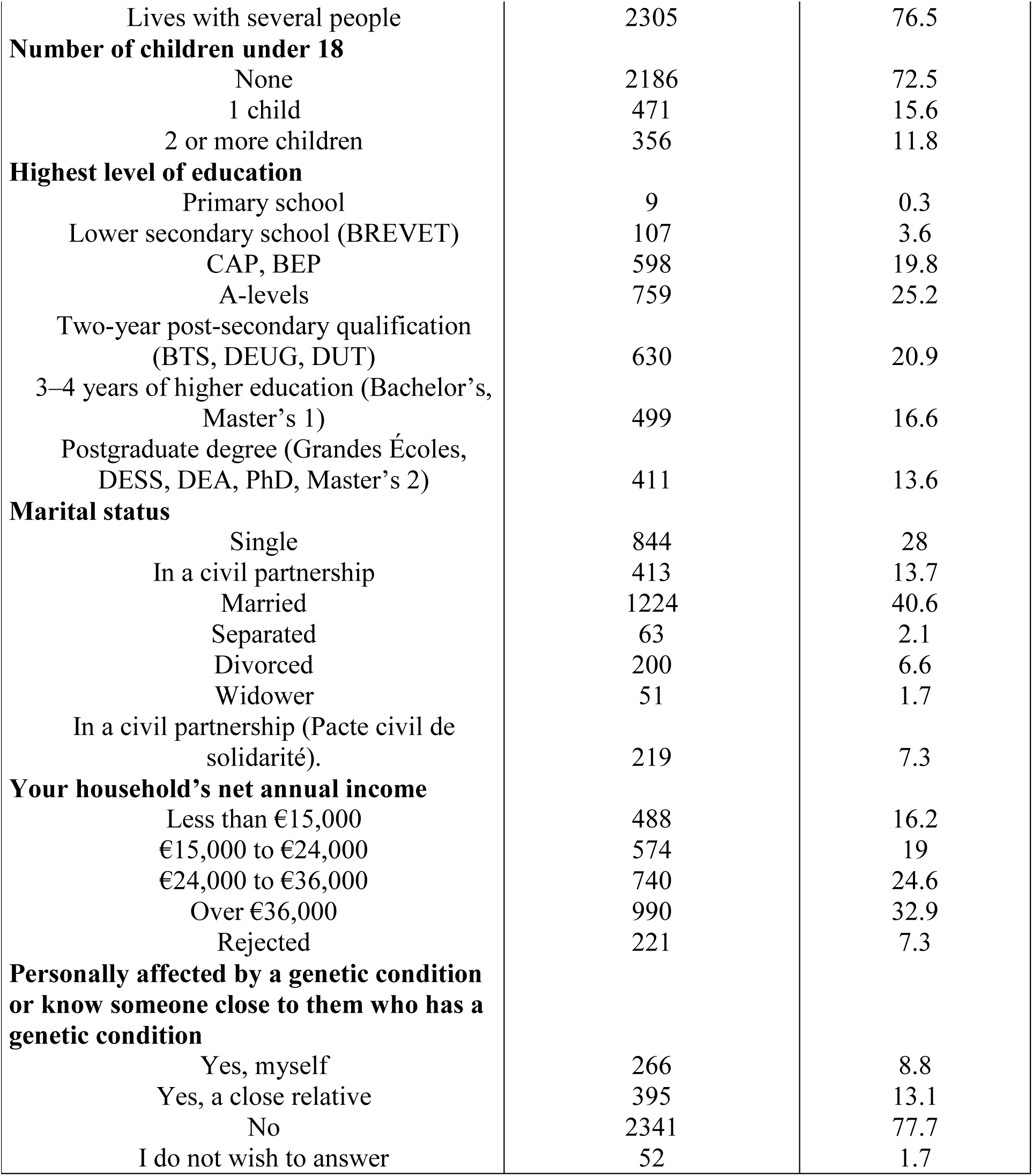
Socio-demographic characteristics of participants.

| Table 1 : Socio-demographic characteristics of participants |  | Number of participants<br>(adjusted figures) | Adjusted % |
| --- | --- | --- | --- |
| **Gender** |  |  |  |
|  | Male | 1435 | 47.6 |
|  | Women | 1578 | 52.4 |
| **Age in years** |  |  |  |
|  | [18–24] | 300 | 10 |
|  | [25–34] | 459 | 15.2 |
|  | [35–44] | 490 | 16.3 |
|  | [45–54] | 523 | 17.4 |
|  | 55 and over | 1240 | 41.2 |
| **Region** |  |  |  |
|  | Paris region | 558 | 18.5 |
|  | North West | 699 | 23.2 |
|  | North East | 663 | 22 |
|  | South West | 318 | 10.6 |
|  | South East | 774 | 25.7 |
| **Type of settlement** |  |  |  |
|  | Rural | 642 | 21.3 |
|  | Fewer than 20,000 inhabitants | 544 | 18.1 |
|  | 20,000 to 100,000 inhabitants | 416 | 13.8 |
|  | 100,000 inhabitants and over | 916 | 30.4 |
|  | Paris | 495 | 16.4 |
| **Current employment status** |  |  |  |
|  | Full-time employee | 1353 | 44.9 |
|  | Part-time employee | 200 | 6.6 |
|  | Self-employed | 109 | 3.6 |
|  | Not currently working, but looking for work | 149 | 4.9 |
|  | Not currently working and not looking for work / unable to work | 105 | 3.5 |
|  | Male/female homemaker | 88 | 2.9 |
|  | Retired | 846 | 28.1 |
|  | Student | 164 | 5.4 |
| **Occupation and socio-professional category of the individual** |  |  |  |
|  | Farmers | 25 | 0.8 |
|  | Self-employed | 113 | 3.8 |
|  | Senior managers | 311 | 10.3 |
|  | Middle-level professionals | 457 | 15.2 |
|  | Employees | 510 | 16.9 |
|  | Workers | 380 | 12.6 |
|  | Pensioners | 846 | 28.1 |
|  | Not in employment | 371 | 12.3 |
| **Number of people in the household** |  |  |  |
|  | Lives alone | 708 | 23.5 |
|  | Lives as a couple | 1113 | 37 |
|  | Lives with 3 or more people | 1192 | 39.6 |

|  |  |  |
| --- | --- | --- |
| Lives with several people | 2305 | 76.5 |
| **Number of children under 18** |  |  |
| None | 2186 | 72.5 |
| 1 child | 471 | 15.6 |
| 2 or more children | 356 | 11.8 |
| **Highest level of education** |  |  |
| Primary school | 9 | 0.3 |
| Lower secondary school (BREVET) | 107 | 3.6 |
| CAP, BEP | 598 | 19.8 |
| A-levels | 759 | 25.2 |
| Two-year post-secondary qualification<br>(BTS, DEUG, DUT) | 630 | 20.9 |
| 3–4 years of higher education (Bachelor's,<br>Master's 1) | 499 | 16.6 |
| Postgraduate degree (Grandes Écoles,<br>DESS, DEA, PhD, Master's 2) | 411 | 13.6 |
| **Marital status** |  |  |
| Single | 844 | 28 |
| In a civil partnership | 413 | 13.7 |
| Married | 1224 | 40.6 |
| Separated | 63 | 2.1 |
| Divorced | 200 | 6.6 |
| Widower | 51 | 1.7 |
| In a civil partnership (Pacte civil de<br>solidarité). | 219 | 7.3 |
| **Your household's net annual income** |  |  |
| Less than €15,000 | 488 | 16.2 |
| €15,000 to €24,000 | 574 | 19 |
| €24,000 to €36,000 | 740 | 24.6 |
| Over €36,000 | 990 | 32.9 |
| Rejected | 221 | 7.3 |
| **Personally affected by a genetic condition**<br>**or know someone close to them who has a**<br>**genetic condition** |  |  |
| Yes, myself | 266 | 8.8 |
| Yes, a close relative | 395 | 13.1 |
| No | 2341 | 77.7 |
| I do not wish to answer | 52 | 1.7 |

All data were compiled into a single database with no individual-level identification. Data quality was ensured through rigorous validation procedures, including automated checks for response time (with exclusion of abnormally fast completions), detection of patterned or inconsistent responses, and targeted review of suspicious response profiles.

Prior to analysis, data cleaning included consistency checks, validation of quantitative variable ranges, verification of filters and respondent bases, assessment of completion rates, management of missing data and identification of coding errors. Invalid observations were excluded.

Descriptive and cross-tabulated analyses were conducted to identify potential inconsistencies. After data cleaning, statistical weighting was applied to correct minor deviations from target quotas and ensure representativeness.

### Variables and data processing

In addition to the standard sociodemographic variables used for quota sampling and weighting, derived variables were constructed from questionnaire responses for analytical purposes. Statistical analyses were performed using COSI software.

Results were exported into structured Excel tables presenting all items, subgroups, and derived indicators (Supplemental Table S1). Outputs included significance testing at the 95% confidence level (with statistically significant differences highlighted), as well as means, standard deviations, and unweighted sample sizes. Percentages were reported as weighted proportions; unweighted bases were used for significance testing across subgroups.

## RESULTS

A total of 3,013 individuals representative of the French population completed an 18-item questionnaire covering four domains: (i) Knowledge of medical genetics; (ii) Perceptions and trust; (iii) Medically indicated genetic testing; and (iv) DTC genetic testing. The main findings are summarized below, with detailed results presented in Figures 1 to 4.

### Knowledge of medical genetics: substantial interest but incomplete understanding (Fig.1)

Overall, 69% of respondents reported an interest in genetics, including 50% who expressed moderate interest whereas only 5% reported no interest.

**Figure 1:**
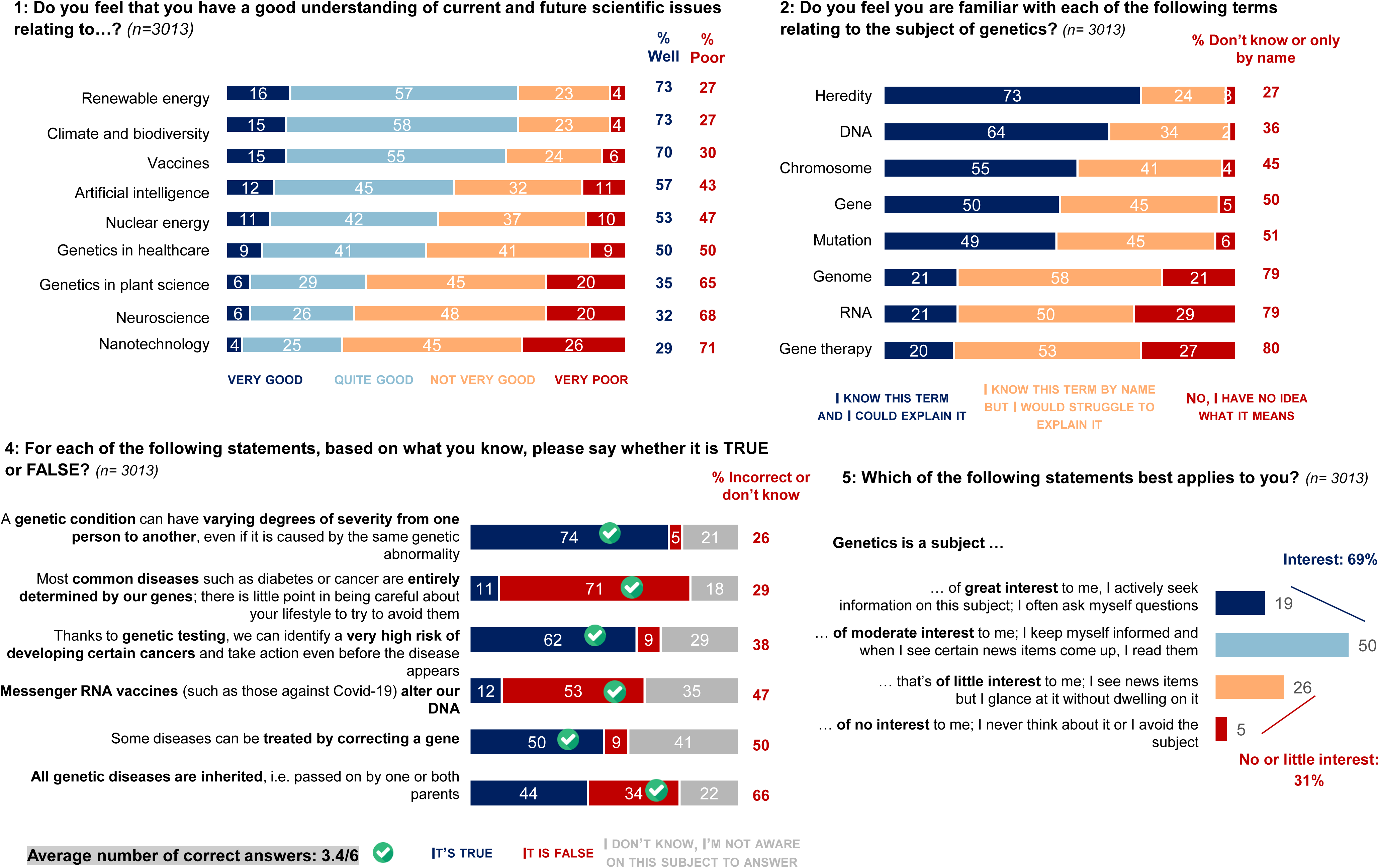
French people’s interest in and knowledge of genetics. The responses to questions 1, 2, 4, and 5 are shown along with the percentage of responses for each question from all respondents (n=3,013). The full questionnaire is available in the Supplementary Data (S1 in French, S2 in English).

However, genetics was ranked among the least well-understood scientific domains, in both healthcare and plant science contexts, alongside artificial intelligence and nuclear science. Approximately half of respondents considered that they understood the general principles.

A more detailed assessment revealed substantial gaps in lexical knowledge. While the terms ‘heredity’ and ‘DNA’ were generally well understood, around 50% of respondents reported difficulties with terms such as ‘chromosome’, ‘gene’, and ‘mutation’. More specialized concepts including genome, RNA, and gene therapy remained poorly understood.

Regarding applied knowledge, three-quarters of respondents correctly recognized that a genetic disease may vary in severity among individuals carrying the same variant. In contrast, only 50% were aware that mRNA vaccines do not alter DNA. The overall mean score was 3.4 out of 6 correct answers (56.6%), and performance was positively associated with educational level and socio-professional category.

### Perceptions and trust: generally favorable climate (Fig.2)

#### Predominantly positive attitudes

A majority of respondents (68%) reported positive feelings towards genetics, such as curiosity, hope, and trust, whereas 32% expressed mistrust or concern. Between 52% and 72% perceived advances in medical genetics as beneficial for both patient care and research governance. However, 28% thought that current misuse in France could lead to serious future consequences.

**Figure 2:**
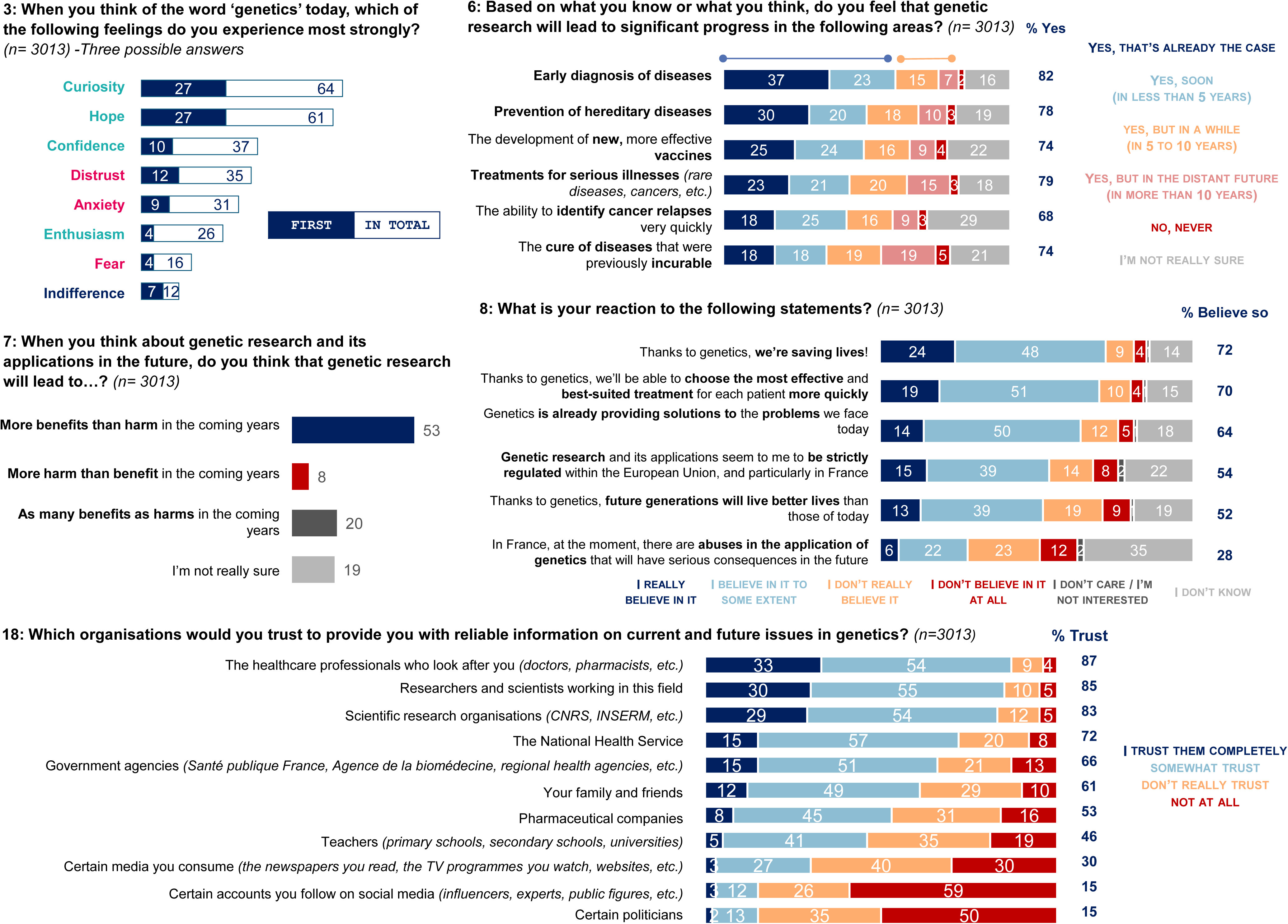
French people’s perceptions of genetics. The responses to questions 3, 6, 7, 8 and 18 are shown along with the percentage of responses for each question from all respondents (n=3,013). The full questionnaire is available in the Supplementary Data (S1 in French, S2 in English).

Half of the respondents anticipated that genetic research would yield more benefits than harms in the coming years, whereas only 8% expected more harm than benefit. These latest proportions were higher among individuals under 35 years of age (12%) and among those affected by a genetic condition (14%).

#### High levels of trust in healthcare professionals

More than 80% of respondents reported trusting healthcare professionals, researchers, and scientific institutions to provide reliable information on genetic issues. This high level of trust highlights the central role of healthcare stakeholders in public communication on genomic medicine.

In contrast, trust in the media, social networks, and political figures was low. Notably, only 46% of respondents reported trusting educators (primary, secondary and higher education), possibly reflecting insufficient training in genetics.

### Medically indicated genetic testing: acceptance and concern (Fig.3)

Eighteen percent of respondents reported that they or a relative had previously been offered a genetic test prescribed by a healthcare professional. While approximately two-thirds correctly understood the concept of genetic predisposition, 34% provided incorrect answers or reported not knowing. In hypothetical clinical scenarios (sudden death in a relative, preconception screening, cancer predisposition, or familial psychiatric disorders), 63–70% of respondents indicated that they would accept genetic testing depending on the indication, while 49% reported that they would accept testing in all proposed scenarios. When asked about receiving a genetic result indicating a high risk of serious disease, 68% of respondents selected proactive or reassuring reactions as their first response: 38% stated that they would prefer to know in order to be monitored earlier, 18% that they would know what to do to reduce their risk, and 12% that they could inform relatives. Conversely, 32% reported feeling anxiety (18%), guilt (9%), or privacy concerns (5%), particularly among individuals under 55 years of age and those affected by a genetic condition. Provided strict legal safeguards are ensured under French law, 77% of respondents indicated willingness to participate in research involving genetic analyses.

**Figure 3:**
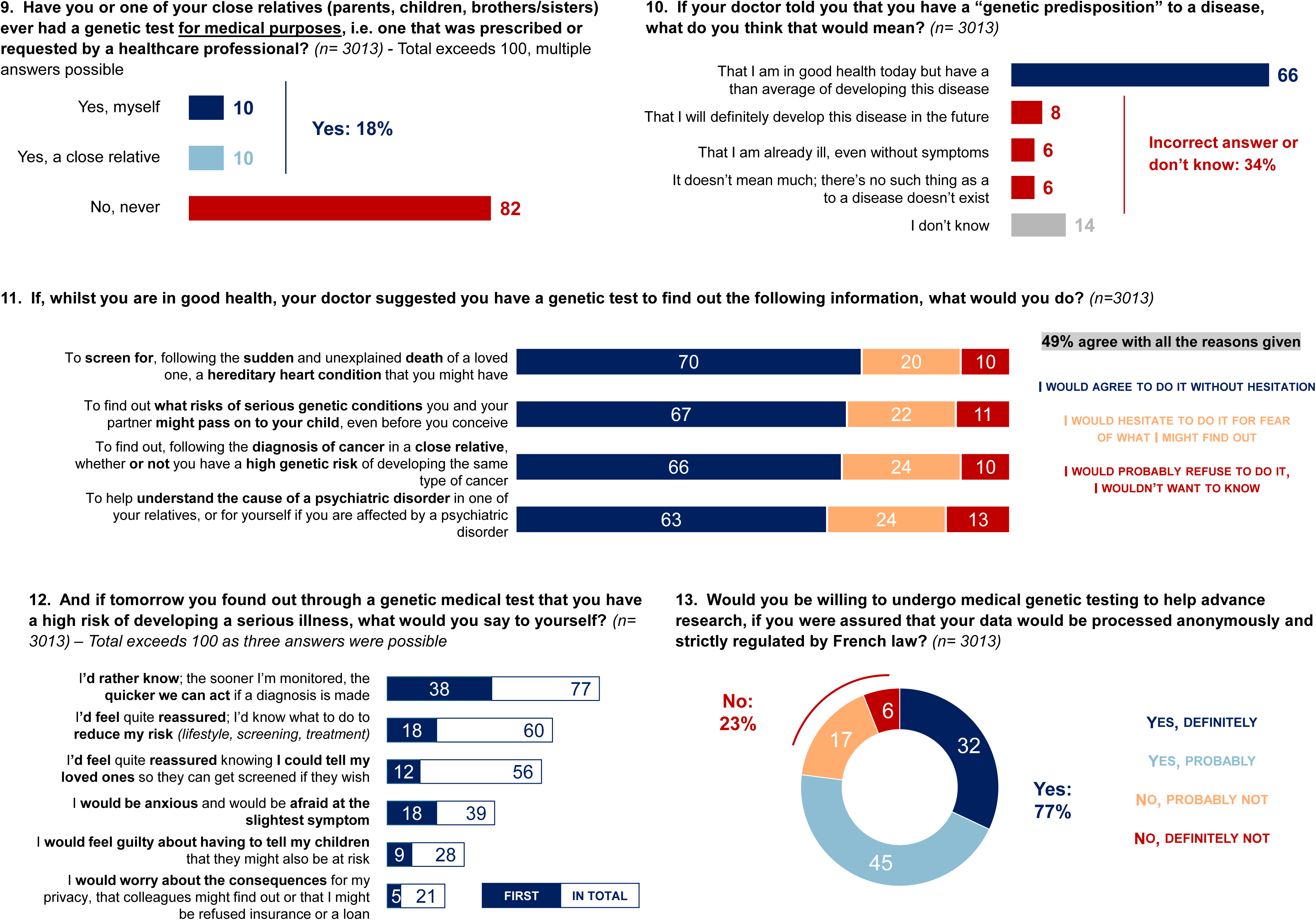
French attitudes towards medical genetic testing. French people’s perceptions of genetics. The responses to questions 9, 10, 11, 12 and 13 are shown along with the percentage of responses for each question from all respondents (n=3,013). The full questionnaire is available in the Supplementary Data (S1 in French, S2 in English).

### Direct-to-consumer genetic testing: an underestimated phenomenon in France (Fig.4)

Despite being prohibited in France, 12% of respondents reported having already ordered a DTC genetic test; 5% for genealogical purposes, 5% for medical purposes, and 2% for both. Among those who had not undergone such testing, 45,3% (n=1206/2661) expressed an interest in doing so. The main motivations were: (i) identifying genetic predispositions to disease, particularly among individuals over 55 years of age; (ii) learning about ethnic and geographical origins; and (iii) personal curiosity.

**Figure 4:**
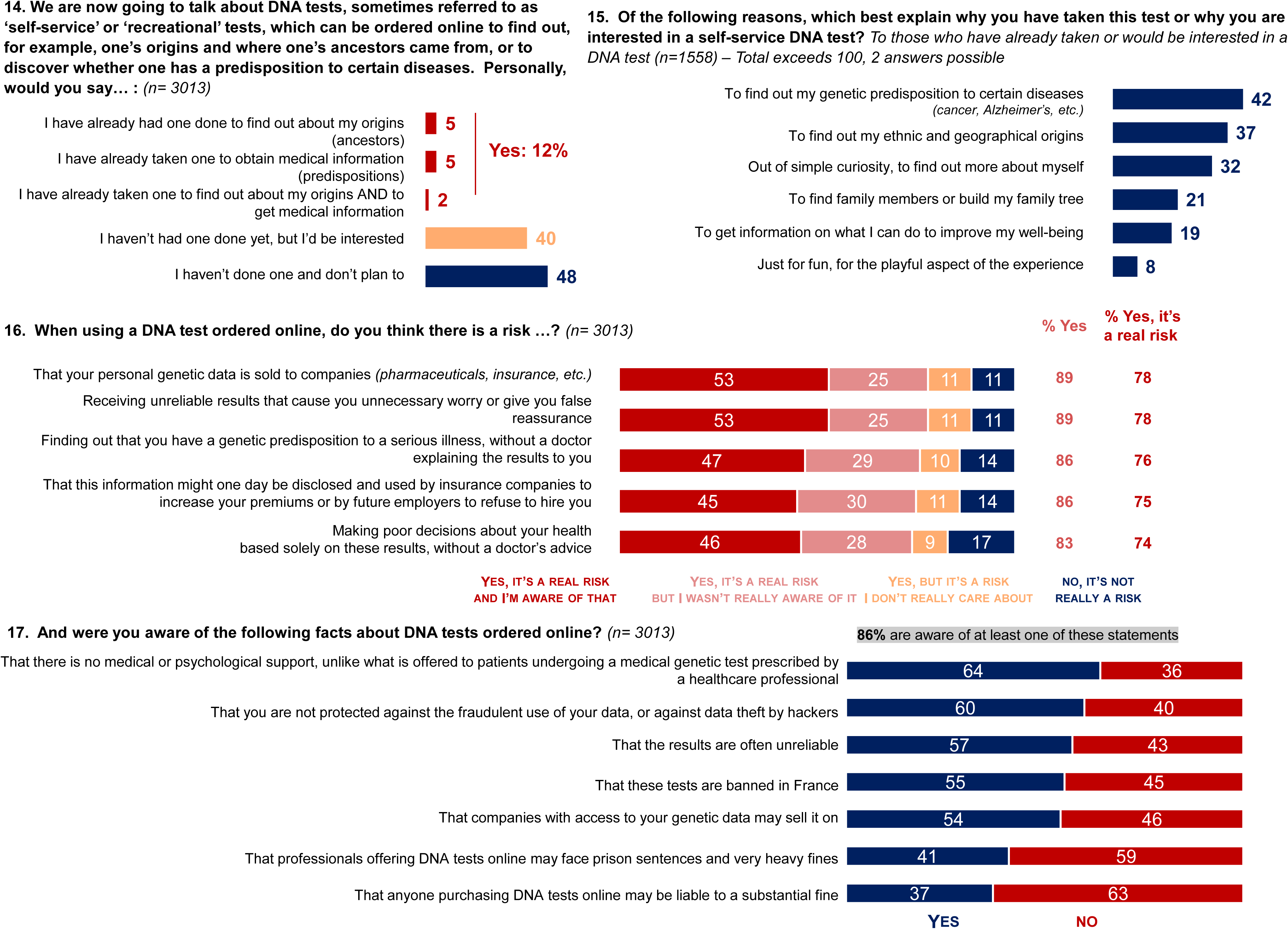
French attitudes towards DTC genetic testing. French people’s perceptions of genetics. The responses to questions 14, 15, 16 and 17 are shown along with the percentage of responses for each question from respondents. The full questionnaire is available in the Supplementary Data (S1 in French, S2 in English).

Regarding associated risks, approximately 75% of respondents acknowledged that ordering such tests online entails real risks. However, only half reported being genuinely aware of these risks. Around one quarter believed there was little or no risk, or did not consider these risks, a proportion that was higher among individuals under 35 years of age and among those who had already undergone testing. Although 86% were aware of at least one potential issue (e.g. lack of legal oversight, absence of medical guidance, or data protection risks), 45% were unaware that these tests are illegal in France.

## DISCUSSION

The survey findings highlight four key findings: (i) Substantial and growing interest in medical genetics among the French population, (ii) Insufficient and fragmented knowledge, (iii) High level of trust in healthcare professionals and scientists, and (iv) Limited awareness of the legal framework and limitations of DTC genetic testing.

### Substantial and growing interest in medical genetics among the French population

A large proportion of respondents (69%) reported an interest in genetics, underscoring the increasing societal relevance of this field. These findings are consistent with the international literature and represent an important foundation for improving public understanding of genetics, enhancing individual autonomy, and supporting informed decision-making. Leveraging this existing interest through culturally adapted communication strategies has been proposed as an effective means of improving genetic counselling practices.(9) In our study, 68% of respondents expressed positive perceptions of genetics, reporting positive feelings. Historically, an Ipsos bva survey conducted in 1988 found that 57% of respondents feared genetic advances, compared to just 16% in the present survey, suggesting increasing societal acceptance over time. This proportion of positive attitudes also appears higher than that reported in the literature. A systematic review of 99 studies published between 2016 and 2022 found that around two-thirds reported generally positive attitudes towards genomics. These perceptions were mainly related to improved understanding of hereditary diseases, identification of genetic causes, and advances in early diagnosis. The development of targeted therapies and personalized medicine was also viewed favorably. However, positive emotional responses specifically related to the psychological implications of genomic testing were less frequently reported in the literature (17.5% of studies) whereas negative attitudes remain common. In our survey, 32% of respondents reported mistrust, concern, or fear. Concerns were primarily related to feelings of guilt and anxiety, as well as potential personal and professional consequences. Similarly, Pearce et al. reported negative emotional responses in 50.9% of studies, including worry, anxiety, stress, fear, and psychological burden.(10) A previous study even reported rates as high as 88%.(11) Furthermore, approximately 44% of motivation studies identified negative perceptions as a barrier to genomic testing. Overall, these findings reflect a dual dynamic combining interest in prevention and anticipation with concerns about the psychological and social implications of genetic information.

In our cohort, 49% of respondents reported willingness to undergo genetic testing across all proposed scenarios (cancer predisposition, risk of sudden death, psychiatric conditions and preconception screening), while approximately 70% expressed willingness for each scenario individually. Regarding preconception genetic screening, a previous French study conducted reported favorable opinion in 91% of participants and willingness in 57%.(12) The relatively homogenous responses observed across different clinical scenarios in our study may reflect limited public understanding, potentially reducing the ability to distinguish between contexts. Middleton et al. showed that attitudes towards genetic testing vary according to knowledge levels, with genetics professionals generally expressing more cautious views.(13) Overall, willingness to undergo testing appears higher in the French population in 2025 than in earlier international studies, in line with increasingly positive perceptions.(10)

### Insufficient and fragmented knowledge

Our findings confirm persistent gaps in genetic knowledge, particularly regarding more complex concepts such as RNA, genomics, and gene therapy. These limitations may compromise the quality of informed consent and the effective exercise of autonomy. As in previous studies, knowledge was positively associated with educational level and socio-professional status. Pearce et al. reported a similar pattern, with high awareness but limited factual understanding, particularly in areas such as genome sequencing, personalized medicine, carrier screening, and pharmacogenomics.(10) Likewise, Haga et al. demonstrated correlations between knowledge and education and Chapman et al., in a large international survey, showed that genetic literacy remains limited even among highly educated individuals.(7,14) Although these data are from 2017, only partial improvements are likely to have occurred since then.

This issue also has an equity dimension. Because knowledge was associated with educational level and socio-professional status, unequal genetic literacy may translate into unequal capacity to benefit from genomic medicine, to question commercial claims, and to avoid misinterpretation of probabilistic results.

### High level of trust in healthcare professionals and scientists

Our results demonstrated high levels of trust in healthcare professionals and researchers, positioning them as central actors in public health communication and awareness campaigns. In contrast, trust in educators, media and political figures was comparatively low. This imbalance highlights the need to strengthen genetics education and communication beyond the healthcare setting. In countries where DTC genetic testing is authorized, the media often constitute a primary source of information.(15) However, the French context differs, as such tests are currently prohibited, limiting public exposure to them outside clinical settings. Media representations of genomics have been described as overly optimistic, potentially generating unrealistic expectations regarding the pace, reliability, and benefits of genomic advances. Improving genetics literacy among journalists and educators therefore appears essential to ensure balanced and accurate dissemination of information.

This finding supports a two-level communication strategy : (i) Healthcare professionals and genetics specialists should remain the reference source for medically relevant information; (ii) Teachers, journalists and public communicators should receive specific training so that basic concepts such as variant, risk, penetrance, carrier status, predisposition and uncertainty are conveyed accurately outside the clinical setting.

Regarding data sharing, Middleton et al. reported substantial international variability but overall limited familiarity with genomic data sharing (64.2%).(16) They also found that perceiving genetic data as separate from other health data was linked to a greater willingness to share such information. In our study, 77% of respondents expressed willingness to participate in research involving genetic testing under the French regulatory framework, compared with just over 50% of the French subset of Middleton’s study conducted in 2020. This higher level of acceptance may reflect strong willingness to contribute to science, as well as confidence in data protection mechanisms such as the European General Data Protection Regulation (GDPR) and national bioethics laws. Similarly, Peyron et al. (2021) showed that the French population values autonomy in accessing genetic information and is generally willing to contribute to research.(17)

### Limited awareness of the legal framework and limitations of DTC genetic testing

Our findings reveal limited awareness of the legal framework governing genetic testing in France. Only 45% of respondents were unaware that DTC is prohibited in France, and many underestimated the limitations of these tests outside a regulated medical context. Notably, 12% of respondents reported having already used DTC genetic testing despite its prohibition, while 40% expressed interest in genealogical and medical purposes, highlighting a gap between public interest, practices, and understanding of legal and ethical implications.

Legal prohibition alone appears insufficient to prevent access to DTC testing or to reduce its attractiveness. Current discussions in France include proposals to authorize DTC testing for genealogical purposes (and not for medical purposes) under a regulated framework. However, such testing raises important concerns, including the validity of inferred ancestry, particularly for underrepresented populations, and the protection of sensitive genetic data often processed outside national jurisdictions. The potential discriminatory use of such data varies across jurisdictions, so continued vigilance is warranted despite existing legal protections in France.

Genealogical testing may also reveal unexpected familial information, such as misattributed parentage or donor conception, with significant psychological and social consequences. In France, access to donor identity for individuals conceived through assisted reproductive technology is regulated by the Commission on Access to Third-Party Donor Data for Individuals Conceived through Assisted Reproductive Technology (CAPADD). Nevertheless, DTC testing may bypass these regulated pathways, raising complex ethical issues regarding consent, privacy, and the rights of all parties involved. More broadly, the absence of structured pre- and post-test counselling challenges the principle of non-maleficence and increases the risk of psychological harm, particularly for vulnerable individuals. On the contrary, medically supervised genetic testing in France requires physician prescription and accredited laboratories within a strict legal framework, ensuring informed consent, analytical validity, data protection, and appropriate disclosure of results. DTC medical testing lacks adequate supervision, increasing the risk of misinterpretation, inappropriate health decisions, and breaches of confidentiality. Moreover, genetic information is inherently familial, meaning that testing may have implications for relatives who have not consented to its disclosure. Reports from clinical practice and the literature highlight the ethical challenges associated with such situations, including unexpected discoveries of non-paternity or intra-familial relationships.(18, 19) Our working group strongly supports maintaining this ban on medical testing, which currently constitutes an illegal medical practice. In this context, the development of a national information strategy on genetic testing appears essential. Such an initiative should aim to help the general public better understand the legal and ethical framework surrounding genetic testing—including its benefits, limitations, and uncertainties—while emphasizing that appropriate medical supervision, psychological support, accreditation, and data protection are essential safeguards.

#### Implications

In the coming years, most individuals in France will likely encounter genetic testing through clinical care, population-based screening, or DTC services. A key finding is that autonomy in genetics extends beyond mere access to testing: it requires understanding what a genetic test can or cannot provide, the probabilistic nature of results, familial implications, and the legal and clinical context. The French population shows interest and openness to medically supervised testing, but the primary challenge is insufficient genetic literacy as access to genetic information grows. Strengthening public autonomy in this area is therefore essential. Genomic acculturation, as described in literature, requires coordinated educational efforts targeting healthcare professionals, students, policymakers and the general public.(9,20) The integration of genetics education into the school curricula has also been emphasized.(21) Improving genetic literacy may enhance critical thinking skills and support decision-making, genetic knowledge has been identified as a major predictor of willingness to undergo genetic testing.(22)

Our findings support three priorities: integrating core genomic concepts into education; developing public-facing information tools led by healthcare professionals and scientific institutions; and ensuring DTC testing policies address not only regulation but also issues related to counseling, data governance, implications for families, and protection against misleading claims.

#### Strengths and limitations

The strengths of this study include its large sample size, national representativeness, and coverage of both medically indicated and DTC genetic testing within the same survey. Several limitations should also be acknowledged. The data are based on self-reported responses collected through an online panel, which may be affected by recall bias, social desirability bias and differential digital access. The cross-sectional design prevents assessment of changes in individual knowledge over time, and the questionnaire measured general understanding rather than in-depth genetic literacy or decision-making in real clinical situations.

In conclusion, medical genetics is entering a phase of widespread societal diffusion. The high level of interest and trust observed in the French population represents a valuable opportunity. However, persistent knowledge gaps, combined with the rapid expansion of DTC genetic testing, create a dual risk: that of nominal autonomy that is not supported by adequate understanding, and that of inequalities in access to information linked to socio-educational factors. In this context, improving genomic literacy is not only an educational challenge but also an ethical imperative essential for safeguarding autonomy, preventing misuse, and ensuring the responsible integration of genomic medicine into the French healthcare system. This survey could be replicated in other European countries to obtain a comprehensive and up-to-date overview of genetics across Europe.

## Data availability statement

All data collected as part of this survey are available in the supplementary information (Supplemental Table S1).

## Author contributions

Conceptualisation and methodology: SM, FP, MMA, PB, ACT, BC, HC, CC, PE, DH, MK, CL, NM, LP, MP, SO, DSL, “Genetics and the General Public” FFGH Ethics Working Group; Funding acquisition: “Genetics and the General Public” FFGH Ethics Working Group; Writing original draft: SM; Writing-review & editing: FP, MMA, PB, ACT, BC, HC, CC, PE, DH, MK, CL, NM, LP, MP, SO, DSL, “Genetics and the General Public” FFGH Ethics Working Group

## Ethical approval

This study was reviewed and approved by the Ethics, Professional Conduct, and Scientific Integrity Committee at the University of Nantes (Comité d’éthique de déontologie et d’intégrité scientifique, CEDIS) n°09122025 (N°IRB: IRB00013074).

## “Genetics and the General Public” FFGH Ethics Working Group

Sandra Mercier^1,2^, François Petit^3^, Micheline Misrahi-Abadou^4^, Philippe Berta^5^, Anne Cambon-Thomsen^6^, Boris Chaumette^7,8^, Hervé Chneiweiss^9^, Célia Crétolle^10^, Patrick Edery^11^, David Heard^12^, Marina Konyukh^13^, Chloé Laeng^14^, Nizar Mahlaoui^15,16^, Laurent Pasquier^17^, Morgane Plutino^18^, Sylvie Odent^17^, Dominique Stoppa-Lyonnet^19^ AND

Katia Andreetti, UMR 7106 - CNRS/ Université Paris II Panthéon-Assas, Paris, France

David Boudeau, President of the French Association of Biology and Geology Teachers, Nantes, France

Estella Castillon, Department of Medical Genetics, Dijon University Hospital, Dijon, France

Philippe Charron, UPMC Univ Paris 6, AP-HP, Hôpital Pitié-Salpêtrière, Centre de Référence Maladies cardiaques héréditaires, Paris, France

Thomas Courtin, Department of Medical Genetics, Necker-Enfants University Hospital, Assistance Publique-Hôpitaux de Paris (AP-HP), Paris, France

Smail Hadj-Rabia, Department of Dermatology, and Reference Centre for Genodermatoses and Rare Skin Diseases (MAGEC), Université Paris Descartes-Paris Cité, INSERM U1163, Institut Imagine, APHP, Hôpital Universitaire Necker-Enfants Malades, 75015 Paris, France

Josiane Jegu, France Assos Santé Association.

Gaëlle Pierron, Service de Génétique, Institut Curie, Paris, France

Virginie Rio, Collectif BAMP !

Damien Sanlaville, Genetics department, GH Est, Hospices Civils de Lyon, Lyon, France

Françoise Shenfield, UCL EGA Institute for Women’s Health, University College Hospital London, London, UK

Christian Siatka, UPR CHROME, Place Gabriel PERI, 30000 Nîmes, France

Stéphane Tirard, Centre François Viète d’épistémologie et d’histoire des sciences et des techniques, Nantes Université, Nantes, France

## Acknowledgments

We would like to extend our sincere thanks to the French Federation of Human Genetics (FFGH) and the coordinators of the ethics working groups, the French Biomedicine Agency, and the Ipsos bva Health team for their support, as well as all 15 partners for co-funding the survey, namely the FHU GenOMedS, the AnDDI-Rares rare disease health network, the SFMPP, the FFGH, as well as the rare disease health networks G2M, NeuroSphinx, SensGene, TeteCou, Cardiogen, Fai2r, Fimarad, MaRIH, MucoCFTR, ORKID, and the Fondamental Foundation.

## Supplementary Information

Table S1: Cross-tabulations based on the complete dataset of questionnaire responses

Document S2: 18-item questionnaire in English

Document S3: 18-item questionnaire in French

