## Supplemental Document S2 for "Knowledge and misconceptions of the French population regarding medical genetics: a survey of 3,000 respondents"

### Questionnaire

### Classification details:

- Age
- Sex
- Socio-professional category
- Level of qualification
- Income level
- Marital status
- Number of children under 18 in the household
- Type of urban area
- Region
- Closeness to the subject of genetics:
    - Presence of a person with a genetic disease among those close to them
    - Presence of someone close who works in the field of genetics

### A) The French public's knowledge of genetics

**Q1: Do you feel you clearly understand the current and future scientific issues concerning…?**

*One answer only per line*

**(2 question units)**

- *Very well*
- *Rather well*
- *Rather poorly*
- *Very poorly*

*Random rotation of items:*

1. Climate and biodiversity
2. Nuclear power
3. Genetics in the field of plants
4. Renewable energy
5. Neuroscience
6. Genetics in the field of health
7. Artificial intelligence
8. Nanotechnology
9. Vaccines

**Q2: Do you feel you know each of the following terms relating to the subject of genetics?**
*One answer only per line*
**(2 question units)**

- *I know this term and could explain it*
- *I know this term by name but I would find it hard to explain*
- *No, I have no idea what it means*

*Random rotation of items:*

1. DNA
2. Chromosome
3. Gene
4. Heredity
5. Mutation
6. Gene therapy
7. Genome
8. RNA

**Q3: When you think of the word "genetics" today, which of the following feelings do you feel most?**
*One answer only per line*
**(2 question units)**

- *Ranked 1st?*
- *Ranked 2nd?*
- *Ranked 3rd?*

*Random rotation of items:*

1. Hope
2. Worry
3. Curiosity
4. Distrust
5. Trust
6. Fear
7. Enthusiasm
8. Indifference

**Q4: For each of the following statements, based on what you know, say whether it is TRUE or FALSE?**
*One answer only per line*
**(2 question units)**

1. It is TRUE
2. It is FALSE
3. I don't know, I am not informed enough on this subject to answer

*Random rotation of items:*

1. Most common diseases such as diabetes or cancer are entirely determined by our genes; it is not much use paying attention to one's lifestyle to try to avoid them (FALSE)
2. All genetic diseases are inherited, i.e. passed on by one or both parents. (FALSE)
3. Messenger RNA vaccines (such as those against Covid-19) modify our DNA (FALSE)
4. Some diseases can be treated by correcting a gene (TRUE)
5. Thanks to a genetic test, one can discover a very high risk of developing certain cancers and act even before the disease appears (TRUE)
6. A genetic disease can have more or less serious impacts from one person to another, even if it is due to the same genetic abnormality. (TRUE)

### B) The French public's interest in genetics

**Q5: Which of the following statements best describes you?**
*One answer only*
**(1 question unit)**
*Random rotation of items:*

**Genetics is a subject …**

1. … of **very strong interest** for me, I actively look for information on this subject, I often ask myself questions
2. … **of moderate interest** for me, I keep informed and when I see certain information go by, I read it
3. … **of low interest** for me, I see information go by but I look at it without dwelling on it.
4. … **of no interest** for me, I never think about it or I avoid the subject.

### C) Genetics and progress

**Q6. Based on what you know or think, do you feel that genetics research will make it possible to achieve significant progress in the following areas?**
*One answer only per line*
**(2 question units)**

- *Yes, it is already the case*
- *Yes, soon (in less than 5 years)*
- *Yes but in some time (in 5 to 10 years)*
- *Yes but in a long time (in more than 10 years)*
- *No, never*
- *I don't really know*

*Random rotation of items:*
1. Early diagnosis of diseases
2. Treatments for serious diseases (rare diseases, cancers, etc.)
3. The prevention of hereditary diseases
4. The ability to identify cancer relapses very quickly
5. The creation of new, more effective vaccines
6. The cure of diseases that were until now incurable

**Q7. When you think about genetics research and its applications in our future, you think that genetics research will produce…?**
*One answer only per line*
**(1 question units)**

*Random rotation of items on the first two items:*
1. More benefits than harms in the coming years
2. More harms than benefits in the coming years
3. As many benefits as harms in the coming years
4. I don't really know

**Q8. What is your reaction to the following statements?**
*One answer only per line*
**(2 question units)**

- *I really believe it*
- *I believe it fairly much*
- *I don't really believe it*
- *I don't believe it at all*
- *I don't care / it doesn't interest me*
- *I don't know*

*Random rotation of items:*

1. Thanks to genetics, future generations will live better than those of today
2. Genetics already provides solutions to the problems we face today
3. Genetics research and its applications seem to me to be strongly regulated within the European Union and particularly in France
4. In France, right now, there are abuses in the applications of genetics that will have serious consequences in the future
5. Thanks to genetics, we save lives!
6. Thanks to genetics, we will be able to choose more quickly the most effective and best-suited treatment for each sick person

### D) Genetics and medical tests: perceptions, knowledge and acceptance

**Definition of a genetic disease: it is a disease caused by one (or more) variation(s) in DNA.**

**Q9. Have you yourself, or someone close to you (parents, children, siblings), ever had a genetic test <u>for medical purposes</u>, i.e. one prescribed or requested by a healthcare professional?**
*Answers 1 and 2 not mutually exclusive*
**(1 question unit)**

1. Yes, myself
2. Yes, someone close to me
3. No, never

**Q10. If your doctor told you that you have a "genetic predisposition" to a disease. What would that mean to you?**
*One answer only*
**(1 question unit)**

*Random rotation of items:*
1. That I am healthy today but have a higher-than-average risk of developing this disease
2. That I am already ill, even without symptoms
3. That I would inevitably develop this disease in the future
4. It doesn't mean much; predisposition to a disease doesn't exist
5. I don't know.

**Q11. If, while you are in good health, your doctor offered you a genetic test to find out the following things, what would you do?**
*One answer only per line*
**(1,5 question unit)**

- *I would agree to do it, without hesitation*
- *I would hesitate to do it for fear of what I might learn*
- *I would probably refuse to do it, I would not want to know*

*Random rotation of items:*

1. To find out which risks of serious genetic diseases you and your partner might pass on to your child, even before conceiving.
2. To find out, after a close relative is diagnosed with cancer, whether or not you have a high genetic risk of developing the same type of cancer
3. To screen, after the sudden and unexplained death of someone close to you, for a hereditary heart condition you might have
4. To help understand the origin of a psychiatric disorder in someone close to you, or for yourself if you are affected by a psychiatric disorder

**Q12. And if tomorrow you learned through a medical genetic test that you have a high risk of developing a serious disease, what would you tell yourself?**
*Three answers possible*
**(2 question units)**

- *Ranked 1st?*
- *Ranked 2nd?*
- *Ranked 3rd?*

*Random rotation of items:*
1. I prefer to know; the earlier I am monitored, the faster we can act if there is a diagnosis.
2. I would be anxious and afraid at the slightest symptom.
3. I would rather feel reassured, I would know what to do to reduce my risk (habits, screenings, treatment).
4. I would feel guilty about having to tell my children that they too could be at risk.
5. I would rather feel reassured to be able to inform those close to me so they can get screened if they wish.
6. I would fear the consequences for my privacy, that colleagues would find out or that I would be refused insurance/credit.

**Q13. Would you be willing to take medical genetic tests to help research advance, if you had the guarantee that your data would be handled anonymously and strictly regulated by French law?**
*One answer only*
**(1 question unit)**

- Yes, certainly
- Yes, probably
- No, probably not
- No, certainly not

### E) Over-the-counter genetic tests: awareness, use and intention, motivations, risk perceptions

**Q14. We will now talk about DNA tests, sometimes called 'self-service' or 'recreational' tests, that can be ordered on the internet to find out, for example, your origins and where your ancestors come from, or to find out whether you have predispositions to certain diseases.**
**Personally, you would say…:**
*One answer only*
**(1 question unit)**

1. I have already taken one to find out my origins (ancestors)
2. I have already taken one to obtain medical information (predispositions)
3. I have already taken one to find out my origins AND to obtain medical information
4. I have not taken one yet, but I would be interested
5. I have not taken one and do not plan to

**If 1 to 4 in Q14**
**Q15. Among the following reasons, which best explain why you took this test or are interested in a self-service DNA test?**
*2 answers possible*
**(1,5 question units)**

1. To find out my ethnic and geographic origins.
2. To find family members or build my family tree.
3. To find out my genetic predispositions to certain diseases (cancer, Alzheimer's, etc.).
4. To obtain information on what I should do to improve my well-being
5. Out of simple curiosity, to learn more about myself.
6. For fun, for the playful side of the experience.

**Q16. When using a DNA test ordered on the internet, do you consider that there is a risk …?**
*One answer only per line*
**(2 question units)**

- *Yes, it is a real risk and I am aware of it*
- *Yes, it is a real risk but I was not really aware of it*
- *Yes, but it is a risk I do not really care about*
- *No, it is not really a risk*

*Random rotation of items:*

1. That your personal genetic data is sold to companies (pharmaceutical, insurance, etc.)
2. Learning that you have a genetic predisposition to a serious disease, without a doctor explaining the results to you
3. Receiving unreliable results that worry you for nothing or wrongly reassure you
4. That this information could one day be disclosed and used by insurers to raise your premiums or by future employers to refuse to hire you
5. Making bad decisions for your health based solely on these results, without a doctor's advice

**Q17. And did you know the following things about DNA tests ordered on the internet?**
*One answer only per line*
*NB: all the options are true*
**(2 question units)**

- *Yes*
- *No*

*Random rotation of items:*

1. That these tests are banned in France
2. That the buyer of DNA tests on the internet can face a substantial fine
3. That the professionals who offer DNA tests on the internet can face prison sentences and very heavy fines
4. That the companies with access to your genetic data are able to resell it
5. That the results are often unreliable
6. That there is no medical or psychological support, unlike what is offered to patients who take a medical genetic test prescribed by a healthcare professional
7. That you are not protected against fraudulent use that could be made of your data, or against data theft by hackers

**Q18. Which of the following would you trust to give you reliable information about the current and future issues around genetics?**
*One answer only per line*
**(2 question units)**

- *Completely trust*
- *Rather trust*

- *Rather do not trust*
- *Do not trust at all*

*Random rotation of items:*

1. Scientific research organisations (CNRS, INSERM, etc.)
2. Researchers, scientists working in this field
3. Your family and friends, those close to you
4. Pharmaceutical companies
5. Teachers (school, high school, universities)
6. Certain media you consult (the newspapers you read, the TV programmes you watch, websites, etc.)
7. Certain accounts you follow on social media (influencers, experts, public figures…)
8. Certain political figures
9. The healthcare professionals who care for you (doctors, pharmacist, etc.)
10. The national health insurance (Assurance maladie)
11. Government agencies (Santé publique France, Agence de la biomédecine, Agences régionales de santé…)

### F) Specific questions for the "pilot" phase

**QP1. How do you perceive the length of the questionnaire?**
*One answer only*

- Too short
- Too long
- Neither too short nor too long

**QP2. Would you say that this questionnaire was …?**
*One answer only per line*

- *Yes, completely*
- *Yes, rather*
- *No, rather not*
- *No, not at all*

1. Easy to understand
2. Easy to answer

**QP3. Did you encounter any difficulties in answering the survey?**
*One answer only*

1. Yes, a lot
2. Yes, a little
3. No

**QP4. And in detail, did you encounter any difficulties in answering the different parts of the survey?**
*One answer only per item*

- *Yes*
- *No*

1. Section on your knowledge of genetics and your interest in genetics
2. Section on your perception of progress in genetics
3. Section on medical tests
4. Section on over-the-counter genetic tests

*To those who encountered difficulties*
**QP5. You said that you encountered difficulties in answering this questionnaire. On which aspects did you encounter difficulties?**
*Several answers possible*

1. Understanding the questions (words used)
2. Clarity of the instructions
3. Length of the questionnaire
4. Length of the lists

5. The True / False quizzes
6. Question format (multiple choice, open-ended questions, etc.)
7. Relevance of the questions to your experience
8. The subject in general
9. Mental fatigue or boredom during the questionnaire
10. Time needed to complete the questionnaire
11. Technical or access problems with the questionnaire
12. Other reasons (please specify in the field below)
