## Supplemental Document S3 for "Knowledge and misconceptions of the French population regarding medical genetics: a survey of 3,000 respondents"

### Questionnaire

### Renseignements signalétiques :

- Age
- Sexe
- Catégorie socioprofessionnelle
- Niveau de diplôme
- Niveau de revenus
- Statut marital
- Nombre d'enfants de moins de 18 au sein du foyer
- Catégorie d'agglomération
- Région
- Proximité avec le sujet de la génétique :
    - Présence de personne atteinte d'une maladie génétique dans son entourage
    - Présence d'un proche qui travaille dans le domaine de la génétique dans son entourage

### A) Les connaissances des Français sur la génétique

**Q1 : Avez-vous le sentiment de bien comprendre les enjeux scientifiques actuels et pour le futur en ce qui concerne…?**
*Une seule réponse possible par ligne*
**(2 unités questions)**

- *Très bien*
- *Plutôt bien*
- *Plutôt mal*
- *Très mal*

*Rotation aléatoire des items :*

1. Le climat et la biodiversité
2. Le nucléaire
3. La génétique dans le domaine des plantes
4. Les énergies renouvelables
5. Les neurosciences
6. La génétique dans le domaine de la santé
7. L'intelligence artificielle
8. Les nanotechnologies
9. Les vaccins

**Q2 : Avez-vous le sentiment de connaître chacun des termes suivants et qui concerne le sujet de la génétique ?**
*Une seule réponse possible par ligne*
**(2 unités questions)**

- *Je connais ce terme et je pourrai l'expliquer*
- *Je connais ce terme de nom mais j'aurai du mal à l'expliquer*
- *Non, je ne sais pas du tout ce qu'il signifie*

*Rotation aléatoire des items :*

1. ADN
2. Chromosome
3. Gène
4. Hérédité
5. Mutation
6. Thérapie génique
7. Génome
8. ARN

**Q3 : Lorsque vous pensez au mot « génétique » aujourd'hui, quels sont les sentiments que vous ressentez le plus parmi les suivants ?**
*Une seule réponse possible par ligne*
**(2 unités questions)**

- *En $1^{er}$ ?*
- *En $2^{nd}$ ?*
- *En $3^{ème}$ ?*

*Rotation aléatoire des items :*

1. Espoir
2. Inquiétude
3. Curiosité
4. Méfiance
5. Confiance
6. Peur
7. Enthousiasme
8. Indifférence

**Q4 : Pour chacune des affirmations suivantes, d'après ce que vous en savez, dites si elle est VRAIE ou FAUSSE ?**
*Une seule réponse possible par ligne*
**(2 unités questions)**

1. C'est VRAI
2. C'est FAUX
3. Je ne sais pas, je ne suis pas assez informé sur ce sujet pour répondre

*Rotation aléatoire des items :*

1. La plupart des maladies courantes comme le diabète ou le cancer sont entièrement déterminées par nos gènes, cela ne sert pas à grand-chose de faire attention à son hygiène de vie pour essayer de les éviter (FAUX)
2. Toutes les maladies génétiques sont héritées, c'est à dire transmises par l'un des parents ou les deux. (FAUX)
3. Les vaccins à ARN messager (comme ceux contre le Covid-19) modifient notre ADN (FAUX)
4. Certaines maladies peuvent être traitées en corrigeant un gène (VRAI)
5. Grâce à un test génétique, on peut découvrir un risque très élevé de développer certains cancers et agir avant même que la maladie n'apparaisse (VRAI)
6. Une maladie génétique peut avoir des impacts plus ou moins graves d'une personne à l'autre, même si elle est due à la même anomalie génétique. (VRAI)

### B) L'intérêt des Français pour la génétique

**Q5 : Laquelle des propositions suivantes vous correspond le mieux ?**
*Une seule réponse possible*
**(1 unité question)**
*Rotation aléatoire des items :*

**La génétique c'est un sujet …**

1. … de **très fort intérêt** pour moi, je cherche activement des informations sur ce sujet, je me pose souvent des questions
2. … **d'intérêt modéré** pour moi, je m'informe et quand je vois certaines informations passer, je les lis
3. … **d'intérêt faible** pour moi, je vois passer des infos mais je les regarde sans m'y attarder.
4. … **sans intérêt** pour moi, je n'y pense jamais ou j'évite le sujet.

### C) Génétique et progrès

**Q6. D'après ce que vous en savez ou ce que vous pensez, avez-vous le sentiment que la recherche en génétique va permettre de réaliser des progrès importants dans les domaines suivants ?**
*Une seule réponse possible par ligne*
**(2 unités questions)**

- *Oui, c'est déjà le cas*
- *Oui, bientôt (dans moins de 5 ans)*
- *Oui mais dans un certain temps (dans 5 à 10 ans)*
- *Oui mais dans longtemps (dans plus de 10 ans)*
- *Non, jamais*
- *Je ne sais pas vraiment*

*Rotation aléatoire des items :*
1. Le diagnostic précoce des maladies
2. Les traitements contre des maladies graves (maladies rares, cancers, etc.)
3. La prévention des maladies héréditaires
4. La capacité à identifier les rechutes de cancer très rapidement
5. La création de nouveaux vaccins plus efficaces
6. La guérison de maladies qui étaient incurables jusque-là

**Q7. Lorsque vous pensez à la recherche en génétique et à ses applications dans notre futur, vous vous dites que la recherche en génétique produira… ?**
*Une seule réponse possible par ligne*
**(1 unité questions)**

*Rotation aléatoire des items sur les deux premiers items :*
1. Plus de bienfaits que de dommages dans les prochaines années
2. Plus de dommages que de bienfaits dans les prochaines années
3. Autant de bienfaits que de dommages dans les prochaines années
4. Je ne sais pas vraiment

**Q8. Quelle est votre réaction face aux affirmations suivantes ?**
*Une seule réponse possible par ligne*
**(2 unités questions)**

- *J'y crois vraiment*
- *J'y crois assez*
- *Je n'y crois pas vraiment*
- *Je n'y crois pas du tout*
- *Je m'en fiche / ça ne m'intéresse pas*
- *Je ne sais pas*

*Rotation aléatoire des items :*

1. Grâce à la génétique, les générations du futur vivront mieux que celles d'aujourd'hui
2. La génétique apporte d'ores et déjà des solutions aux problèmes que nous rencontrons aujourd'hui
3. La recherche en génétique et ses applications me semblent fortement encadrées par la réglementation au sein de l'Union Européenne et notamment en France
4. En France, en ce moment, il y a des dérives dans les applications de la génétique qui auront des conséquences graves dans le futur
5. Grâce à la génétique, on sauve des vies !
6. Grâce à la génétique, on pourra choisir plus vite le traitement le plus efficace et le plus adapté à chaque personne malade

### D) Génétique et tests médicaux : perceptions, connaissances et adhésion

**Définition d'une maladie génétique : il s'agit d'une maladie causée par une (ou plusieurs) variation(s) de l'ADN.**

**Q9. Vous-même ou l'un de vos proches (parents, enfants, frères/sœurs), avez-vous déjà réalisé un test génétique à visée médicale, c'est-à-dire qui a été prescrit ou demandé par un professionnel de santé ?**
*Réponses 1 et 2 non exclusives*
**(1 unité question)**

1. Oui, moi-même
2. Oui, l'un de mes proches
3. Non, jamais

**Q10. Si votre médecin vous informait que vous avez une "prédisposition génétique" à une maladie. Qu'est-ce que cela voudrait dire selon vous ?**
*Une seule réponse possible*
**(1 unité question)**

*Rotation aléatoire des items :*
1. Que je suis en bonne santé aujourd'hui mais que j'ai un risque plus élevé que la moyenne de développer cette maladie
2. Que je suis déjà malade, même sans symptôme
3. Que je développerais forcément cette maladie à l'avenir
4. Cela ne veut pas dire grand-chose, la prédisposition à une maladie ça n'existe pas
5. Je ne sais pas.

**Q11. Si, alors que vous êtes en bonne santé, votre médecin vous proposait de réaliser un test génétique pour savoir les choses suivantes, que feriez-vous ?**
*Une seule réponse possible par ligne*
**(1,5 unité question)**

- *J'accepterais de le faire, sans hésiter*
- *J'hésiterais à le faire par crainte de ce que je pourrais apprendre*
- *Je refuserais probablement de le faire, je ne voudrais pas savoir*

*Rotation aléatoire des items :*

1. Pour savoir quels risques de maladies génétiques graves, vous et votre conjoint, risquez de transmettre à votre enfant, avant même de le concevoir.
2. Pour savoir, après la découverte d'un cancer chez un parent proche, si vous avez un risque génétique élevé ou pas de développer le même type de cancer
3. Pour dépister, après un décès soudain et inexpliqué d'un proche, une anomalie cardiaque héréditaire dont vous pourriez souffrir
4. Pour aider à comprendre l'origine d'un trouble psychiatrique chez l'un de vos proches ou pour vous-même si vous êtes concerné par un trouble psychiatrique

**Q12. Et si demain vous appreniez grâce à un test médical génétique que vous avez un risque important de développer une maladie grave, qu'est-ce que vous vous diriez ?**
*Trois réponses possibles*
**(2 unités question)**

- *En 1^er ?*
- *En 2^ème ?*
- *En 3^ème ?*

*Rotation aléatoire des items :*
1. Je préfère le savoir, plus je serais suivi(e) tôt et plus on pourra agir vite en cas de diagnostic.
2. Je serais angoissé(e) et j'aurais peur au moindre symptôme.
3. Je serais plutôt rassuré, je saurais quoi faire pour réduire mon risque (habitudes, dépistages, traitement).
4. Je me sentirais coupable de devoir dire à mes enfants qu'ils pourraient aussi être à risque.
5. Je serais plutôt rassuré de pouvoir informer mes proches pour qu'ils se dépistent s'ils le souhaitent.
6. Je craindrais les conséquences pour ma vie privée, que des collègues l'apprennent ou qu'on me refuse une assurance/crédit.

**Q13. Seriez-vous prêt(e) à réaliser des tests génétiques médicaux pour aider la recherche à avancer, si vous aviez la garantie que vos données seront traitées de façon anonyme et strictement encadrées par la législation française ?**
*Une seule réponse possible*
**(1 unité question)**

- Oui, certainement
- Oui, probablement
- Non, probablement pas
- Non, certainement pas

### E) Tests génétiques en accès libre : notoriété, utilisation et intention, motivations, perceptions des risques

**Q14. Nous allons parler maintenant des tests ADN parfois appelés tests 'en libre-service' ou 'récréatifs' que l'on peut commander sur internet pour connaître par exemple ses origines et savoir d'où proviennent ses ancêtres ou encore pour savoir si on a des prédispositions pour certaines maladies.**
**Personnellement, vous diriez… :**
*Une seule réponse possible*
**(1 unité question)**

1. J'en ai déjà fait un pour connaitre mes origines (ancêtres)
2. J'en ai déjà fait un pour avoir des informations médicales (prédispositions)
3. J'en ai déjà fait un pour connaitre mes origines ET avoir des informations médicales
4. Je n'en ai pas encore fait, mais cela m'intéresserait
5. Je n'en ai pas fait et je n'envisage pas de le faire

**Si 1 à 4 en Q14**
**Q15. Parmi les raisons suivantes, quelles sont celles qui expliquent le plus que vous ayez fait ce test ou que vous soyez intéressé(e) par un test ADN en libre-service ?**
*2 réponses possibles*
**(1,5 unités question)**

1. Pour connaître mes origines ethniques et géographiques.
2. Pour retrouver des membres de ma famille ou construire mon arbre généalogique.
3. Pour connaître mes prédispositions génétiques à certaines maladies (cancer, Alzheimer, etc.).
4. Pour obtenir des informations sur ce que je dois faire pour améliorer mon bien-être
5. Par simple curiosité, pour en savoir plus sur moi-même.
6. Par jeu, pour le côté ludique de l'expérience.

**Q16. Quand on utilise un test ADN que l'on commande sur internet, estimez-vous qu'il y a un risque … ?**
*Une seule réponse possible par ligne*
**(2 unités question)**

- *Oui c'est un risque réel et j'en ai conscience*
- *Oui c'est un risque réel mais je n'en avais pas vraiment conscience*
- *Oui mais c'est un risque dont je me fiche un peu*
- *Non, ce n'est pas vraiment un risque*

*Rotation aléatoire des items :*
1. Que vos données génétiques personnelles soient vendues à des entreprises (pharmaceutiques, assurances, etc.)
2. D'apprendre que l'on a une prédisposition génétique à une maladie grave, sans qu'un médecin ne vous explique les résultats
3. De recevoir des résultats peu fiables qui vous inquiètent pour rien ou vous rassurent à tort
4. De voir que ces informations soient un jour divulguées et utilisées par des assurances pour augmenter vos cotisations ou par de futurs employeurs pour refuser une embauche
5. De prendre de mauvaises décisions pour votre santé en vous basant uniquement sur ces résultats, sans l'avis d'un médecin

**Q17. Et saviez-vous les choses suivantes à propos des tests ADN que l'on commande sur internet ?**
*Une seule réponse possible par ligne*
*NB : toutes les modalités sont vraies*
**(2 unités question)**

- *Oui*
- *Non*

*Rotation aléatoire des items :*
1. Que ces tests sont interdits en France
2. Que l'acheteur de tests ADN sur internet peut être puni d'une amende importante
3. Que les professionnels qui proposent des tests ADN sur internet peuvent être punis de peines d'emprisonnement et à de très fortes amendes
4. Que les entreprises qui ont accès à vos données génétiques ont la possibilité de les revendre
5. Que les résultats ne sont souvent pas fiables
6. Qu'il n'y a pas d'accompagnement médical, ni psychologique, contrairement à ce qui est proposé aux patients qui passent un test génétique médical prescrit par un professionnel de santé
7. Que vous n'êtes pas protégé contre l'utilisation frauduleuse qui pourrait être faite de vos données, ou contre le vol de données par des pirates informatiques

**Q18. A quels acteurs feriez-vous confiance pour vous apporter une information fiable sur les enjeux actuels et futurs de la génétique ?**
*Une seule réponse possible par ligne*

**(2 unités questions)**

- *Tout à fait confiance*
- *Plutôt confiance*
- *Plutôt pas confiance*
- *Pas du tout confiance*

*Rotation aléatoire des items :*

1. Les organismes de recherche scientifique (CNRS, INSERM, etc.)
2. Les chercheurs, les scientifiques qui travaillent dans ce domaine
3. Vos proches, votre entourage
4. Les laboratoires pharmaceutiques
5. Les enseignants (école, lycée, universités)
6. Certains médias que vous consultez (les journaux que vous lisez, les émissions de télévision que vous regardez, sites internet, etc.)
7. Certains comptes que vous suivez sur les réseaux sociaux (influenceurs, experts, personnalités…)
8. Certaines personnalités politiques
9. Les professionnels de santé qui vous suivent (médecins, pharmacien, etc.)
10. L'Assurance maladie
11. Les agences gouvernementales (Santé publique France, Agence de la biomédecine, Agences régionales de santé…)

### F) Questions spécifiques pour la phase « pilote »

**QP1. Quelle est votre perception de la durée du questionnaire ?**
*Une seule réponse possible*

- Trop courte
- Trop longue
- Ni trop courte, ni trop longue

**QP2. Diriez-vous de ce questionnaire qu'il était … ?**
*Une seule réponse possible par ligne*

- *Oui, tout à fait*
- *Oui, plutôt*
- *Non, plutôt pas*
- *Non, pas du tout*

1. Facile à comprendre
2. Facile à répondre

**QP3. Avez-vous rencontré des difficultés pour répondre à l'enquête ?**
*Une seule réponse possible*

1. Oui, beaucoup
2. Oui, un peu
3. Non

**QP4. Et dans le détail, avez-vous rencontré des difficultés pour répondre aux différentes parties de l'enquête ?**
*Une seule réponse possible par item*

- *Oui*
- *Non*

1. Partie sur vos connaissances de la génétique et votre intérêt sur la génétique
2. Partie sur votre perception des progrès de la génétique
3. Partie sur les tests médicaux
4. Partie sur les tests génétiques en accès libre

*A ceux qui ont rencontré des difficultés*
**QP5. Vous avez dit que vous aviez rencontré des difficultés pour répondre à ce questionnaire. Sur quels aspects avez-vous rencontré des difficultés ?**
*Plusieurs réponses possibles*

1. Compréhension des questions (mots utilisés)
2. Clarté des instructions
3. Longueur du questionnaire
4. Longueur des listes

5. Les quiz Vrai / Faux
6. Format des questions (choix multiples, questions ouvertes, etc.)
7. Pertinence des questions par rapport à votre expérience
8. Le sujet en général
9. Fatigue mentale ou ennui pendant le questionnaire
10. Temps nécessaire pour compléter le questionnaire
11. Problèmes techniques ou d'accès au questionnaire
12. Autres raisons (veuillez préciser dans le champ ci-dessous)
